# Tracking the COVID-19 pandemic in Australia using genomics

**DOI:** 10.1101/2020.05.12.20099929

**Authors:** Torsten Seemann, Courtney R Lane, Norelle L Sherry, Sebastian Duchene, Anders Gonçalves da Silva, Leon Caly, Michelle Sait, Susan A Ballard, Kristy Horan, Mark B Schultz, Tuyet Hoang, Marion Easton, Sally Dougall, Timothy P Stinear, Julian Druce, Mike Catton, Brett Sutton, Annaliese van Diemen, Charles Alpren, Deborah A Williamson, Benjamin P Howden

## Abstract

**BACKGROUND:** Whole-genome sequencing of pathogens can improve resolution of outbreak clusters and define possible transmission networks. We applied high-throughput genome sequencing of SARS-CoV-2 to 75% of cases in the State of Victoria (population 6.24 million) in Australia.

**METHODS:** Cases of SARS-CoV-2 infection were detected through active case finding and contact tracing. A dedicated SARS-CoV-2 multidisciplinary genomic response team was formed to enable rapid integration of epidemiological and genomic data. Phylodynamic analysis was performed to assess the putative impact of social restrictions.

**RESULTS:** Between 25 January and 14 April 2020, 1,333 COVID-19 cases were reported in Victoria, with a peak in late March. After applying internal quality control parameters, 903 samples were included in genomic analyses. Sequenced samples from Australia were representative of the global diversity of SARS-CoV-2, consistent with epidemiological findings of multiple importations and limited onward transmission. In total, 76 distinct genomic clusters were identified; these included large clusters associated with social venues, healthcare facilities and cruise ships. Sequencing of sequential samples from 98 patients revealed minimal intra-patient SARS-CoV-2 genomic diversity. Phylodynamic modelling indicated a significant reduction in the effective viral reproductive number (*R_e_*) from 1.63 to 0.48 after the implementation of travel restrictions and population-level physical distancing.

**CONCLUSIONS:** Our data provide a comprehensive framework for the use of SARS-CoV-2 genomics in public health responses. The application of genomics to rapidly identify SARS-CoV-2 transmission chains will become critically important as social restrictions ease globally. Public health responses to emergent cases must be swift, highly focused and effective.

## INTRODUCTION

The coronavirus disease (COVID-19) pandemic caused by severe acute respiratory syndrome coronavirus 2 (SARS-CoV-2) is a global public health emergency on a scale not witnessed in living memory. First reports in December 2019 described a cluster of patients with pneumonia, linked to a market in Wuhan, China.^1,2^ Subsequent testing revealed the presence of a previously unknown coronavirus, now termed SARS-CoV-2, with the associated disease termed COVID-19.^2^

Initial laboratory responses included early characterization and release of the viral whole genome sequence (strain Wuhan-Hu-1) in early January 2020,^2^ which enabled rapid development of reverse-transcriptase polymerase chain reaction (RT-PCR) diagnostics.^3^ To date, laboratory testing has played a critical role in defining the epidemiology of the disease, informing case and contact management, and reducing viral transmission.^4^ In addition to facilitating the development of diagnostic tests, whole genome sequencing (WGS) can be used to detect phylogenetic clusters of SARS-CoV-2,^5^ with many laboratories now making genomic data publicly available.^6,7^

For other viral pathogens genomic surveillance has been used to detect and respond to putative transmission clusters,^8,9^ and to provide information on the possible source of individual cases.^10^ To ensure maximal public health utility, genomics-informed public health responses require detailed integration of genomic and epidemiological data, which in turn requires close liaison between laboratories and public health agencies. Here, we combined extensive WGS and epidemiologic data to investigate the source of individual cases of COVID-19 in Victoria, Australia. This report describes the key findings from the first 1,333 cases of COVID-19 in our setting, and demonstrates the integration of genomics-based COVID-19 surveillance into public health responses.

## METHODS

### Setting, data sources and COVID-19 genomics response group

In the State of Victoria, Australia (population approximately 6.24 million) all samples positive for SARS-CoV-2 by RT-PCR are forwarded to the Doherty Institute Public Health Laboratories,^11^ for confirmation and genomics analysis. We conducted a retrospective, observational study of all patients in Victoria with confirmed COVID-19 with a diagnosis prior to 14 April 2020, including collection of detailed demographic and risk factor information on each case. Epidemiological clusters were defined as clusters containing three or more cases with a common source exposure (e.g. healthcare facility).

To rapidly implement SARS-CoV-2 genomic analysis into local public health responses, a COVID-19 genomics response team was convened, including representatives from the state health department, virology laboratory, and the public health genomics laboratory (genomic epidemiologist, bioinformaticians and medical microbiologists). Laboratory and bioinformatic workflows were developed to support large-scale rapid processing of samples, enabling genomic sequencing and bioinformatic analysis of 96 samples in an approximately 45-hour time period. The response team held online meetings to enable interactive reporting of genomic epidemiological analyses and facilitate rapid translation of genomic findings into public health responses.

### Genomic sequencing and bioinformatic analysis

Detailed methods are provided in the Supplementary Appendix. In brief, extracted RNA from SARS-CoV-2 RT-PCR positives samples underwent tiled amplicon PCR using either ARTIC version 1 or version 3 primers ^12^ using published protocols,^13^ and Illumina sequencing. Reads were aligned to the reference genome (Wuhan Hu-1; GenBank MN908947.3) and consensus sequences generated. We applied quality control checks on consensus sequences, requiring ≥80% genome recovered, ≤25 single nucleotide polymorphisms (SNPs) from the reference genome, and ≤300 ambiguous or missing bases for sequences to ‘pass’ QC (Supplementary Figure S1). For phylogenetic analysis, a single sequence was selected per patient, and genomic clusters defined as two or more related sequences using Cluster Picker^14^; additionally, recently proposed lineages were also determined.^15^ Intra-patient sequence variability was assessed by comparing different samples from the same patient (Supplementary Appendix).

### Estimation of population parameters using phylodynamic analyses

We conducted Bayesian phylodynamic analyses using the 903 genome samples from Victoria, sampled between 25 January and 14 April 2020 using BEAST2.5 (Supplementary Appendix).^16^ We calibrated the molecular clock using the sample collection times, and inferred epidemiological dynamics assuming a birth-death skyline model including two intervals for *R_e_* estimated in the analysis. We assumed a duration of infection of 9.68 days to match independent epidemiological estimates reported by local mathematical modelling data.^17^

### Data sharing and availability

Consensus sequences and Illumina sequencing reads were deposited into GenBank under BioProject PRJNA613958 (Supplementary Data). Consensus genome sequences are also available from https://github.com/MDU-PHL/COVID19-paper.

### Statistical analysis

Associations between categorical data were made using a chi-squared test, and differences in non-normally distributed numerical data using the Wilcoxon Rank sum test. All statistical analyses were performed using R (v.3.6.3).

### Ethics and study oversight

Data were collected in accordance with the Victorian Public Health and Wellbeing Act 2008. Ethical approval was received from the University of Melbourne Human Research Ethics Committee (study number 1954615.3). All authors vouch for the integrity and completeness of data and analyses.

## RESULTS

### Demographic characteristics of cases

Over the study period, there were 1,333 laboratory-confirmed cases of COVID-19 in Victoria. Of these, 631/1,333 (54.2%) were male, and the median age was 47 years (IQR 29 to 61) (Table 1). The majority of cases (827/1,333, 62.0%) were identified in returning travelers, most commonly from north-west Europe and the Americas, and 360 (27.0%) in known COVID-19 contacts (Table 1). Cases in Victoria peaked in mid-March, then declined over the study period, consistent with population-level public health interventions (Figure 1). In total, 134/1,333 (10.1%) cases were from an unknown source within Australia.

**Table 1:** Demographic and risk factor data for Victorian COVID-19 cases and those with available sequence data, 19 January to 14 April 2020.

| Characteristic | Number (% of total) |
| --- | --- |
| Sex |  |
| Male | 473 (53.6%) |
| Female | 410 (45.4%) |
| Unknown | 20 (2.2%) |
| Median age (years) (IQR) |  |
| All | 46 (29-60) |
| Males | 45 (28-58) |
| Females | 46 (29-46) |
| Healthcare worker | 109 (11.9%) |
| Residence in metropolitan region | 755 (86.5%) |
| Putative source of acquisition |  |
| Overseas travel | 557 (61.7%) |
| Contact with known case | 260 (28.8%) |
| Unknown | 81 (9.0%) |
| Region of travel (for travel-associated cases, n=557)* |  |
| Oceania | 61 (10.9%) |
| North-West Europe | 230 (41.3%) |
| Southern and Eastern Europe | 41 (7.4%) |
| North Africa and the Middle East | 24 (4.3%) |
| South-East Asia | 35 (6.3%) |
| North-East Asia | 12 (2.2%) |
| Southern and Central Asia | 7 (1.3%) |
| Americas | 169 (30.3%) |
| Sub-Saharan Africa | 8 (1.4%) |
| Sample site |  |
| Nasopharyngeal swab / nasal swab | 839 (92.9%) |
| Lower respiratory tract specimen | 13 (1.4%) |
| Unknown / Other | 51 (5.7%) |

\* Cases travelling to more than one region were counted more than once

**Figure 1:**
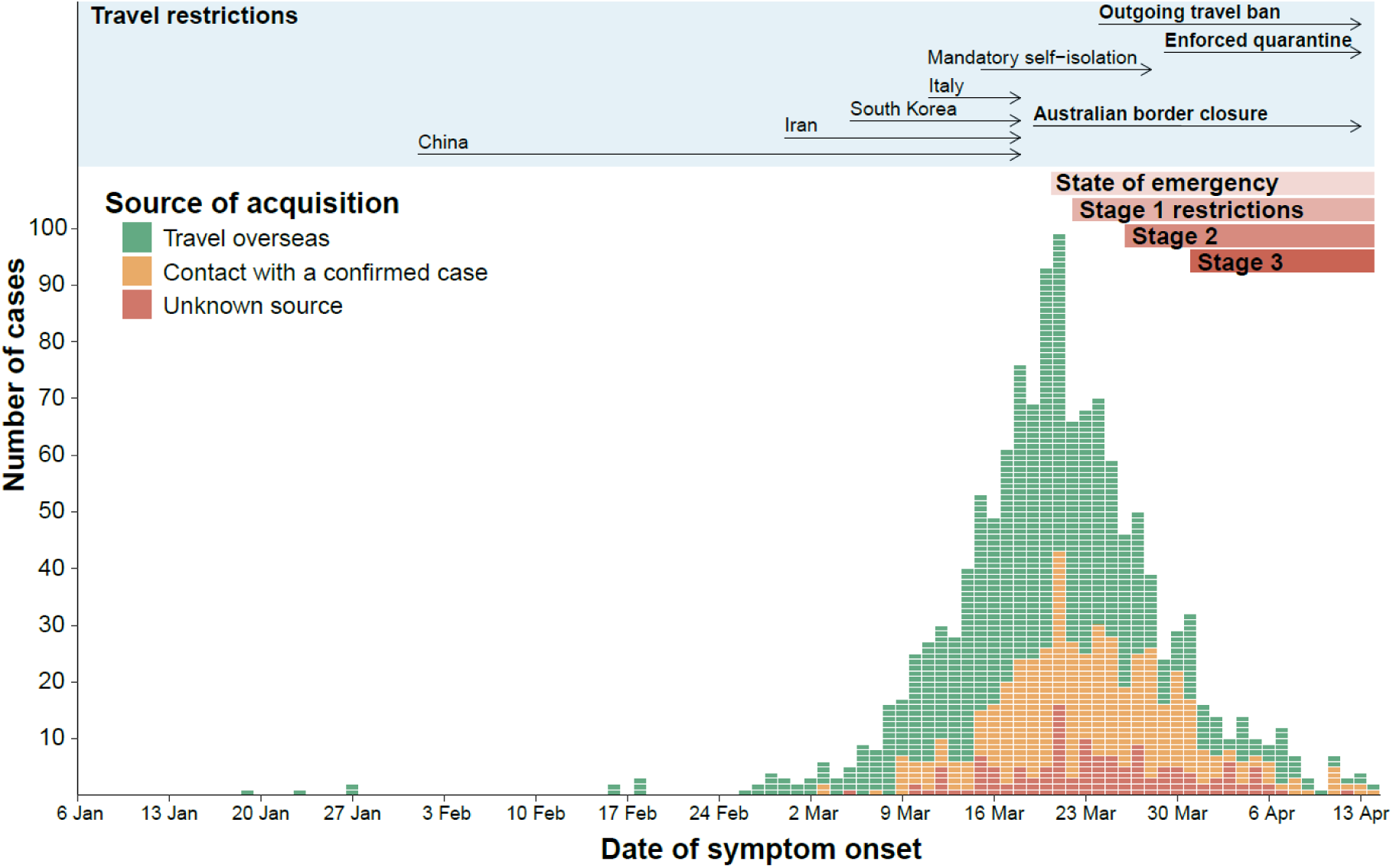
Epidemic curve of COVID-19 cases, by putative mode of acquisition, and implementation of key public health interventions, Victoria, Australia, 06 January – 14 April 2020. Cases were categorized as (i) travel overseas if reporting travel in the 14 days prior to symptom onset or (ii) contact with a confirmed case if no overseas travel reported and case contact occurred within the same time period. Cases are plotted by reported date of symptom onset, or if unknown, date of initial specimen collection. State of emergency declaration introduced a ban on large gatherings and mandatory social distancing of 4m^2^ per person. Stage 1 restrictions introduced a shutdown of nonessential services, followed shortly after by early commencement of school holidays. Stage 2 restrictions expanded shut down of non-essential services, and Stage 3 introduced an enforceable stay-at-home order and limited non-household groups to 2 people.

### High-throughput prospective viral sequencing and genomic epidemiology

A total of 1,242 samples from 1,075 patients were sequenced during the study period, representing 80.7% of all cases (Supplementary Figure S2). There were no significant demographic differences between cases with and cases without included sequence data (Supplementary Table 1). Of the 1,242 samples, 1,085 (87.3%) passed QC; after excluding duplicate patients from cases, 903 samples (68% of cases) were included in the final alignment. Included sequences had a significantly lower PCR cycle threshold (Ct) value than sequences excluded from the final alignment (median 27, IQR 22-31 vs. median 36, IQR 32-38, for excluded sequences; P < 0.001,) (Supplementary Table S2 and Supplementary Figure S3). As reported elsewhere, we found relatively little genetic variation across the genomes, with a maximum of 15 SNPs observed relative to the Wuhan-1 reference (median 7 SNPs, IQR 6-9).

Almost all second-level lineages from a recently proposed SARS-CoV-2 genomic nomenclature ^15^ were identified in the dataset (excluding lineage A.4), suggesting that Victorian samples were representative of the global diversity of published SARS-CoV-2 sequences, consistent with epidemiological findings (Figure 2 and Table 1).

**Figure 2:**
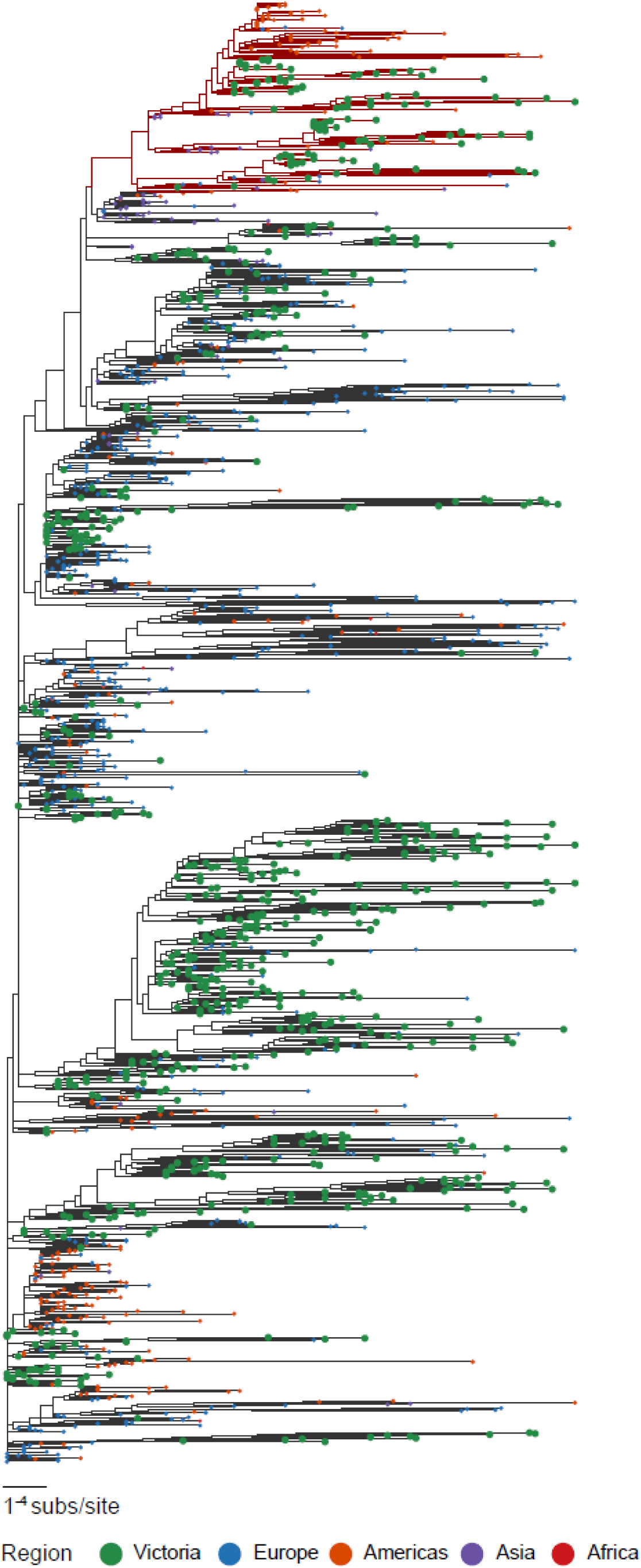
Phylogenetic tree of SARS-CoV-2 sequences from Victorian COVID-19 patients in context of publicly available international SARS-CoV-2 sequences. Maximum likelihood tree of Victorian SARS-CoV-2 sequences and a subset random selection of international sequences representing global genomic diversity, colored by region of origin. Victorian isolates, in green, have been emphasized through increased size, and represent the global diversity of the sampled SARS-CoV-2 population. Branch color represents Pangolin lineage A (red) or B (black).

### Genomic clustering amongst Victorian COVID-19 cases

In total, 737 samples belonged to a genomic cluster, representing 81.6% of the samples in the final dataset. Overall, 76 genomic clusters were identified, with a median of 5 cases per cluster (range 2 to 75, IQR 2-11 cases) and median duration of 13 days (IQR 5-22 days) (Figure 3A), consistent with repeated introduction and limited subsequent local transmission. Of the 76 clusters, 34 (45%) contained only cases reporting overseas travel; a further 34/76 clusters (45%) contained both travel-associated and locally-acquired cases, with the first sampled case reporting overseas travel in 27/34 (79%) of these clusters. There was strong concordance between epidemiological and genomic clusters, with a median of 100% (IQR 85-100%) of cases within an epidemiological cluster contained within a single dominant genomic cluster (Figure 3B).

**Figure 3:**
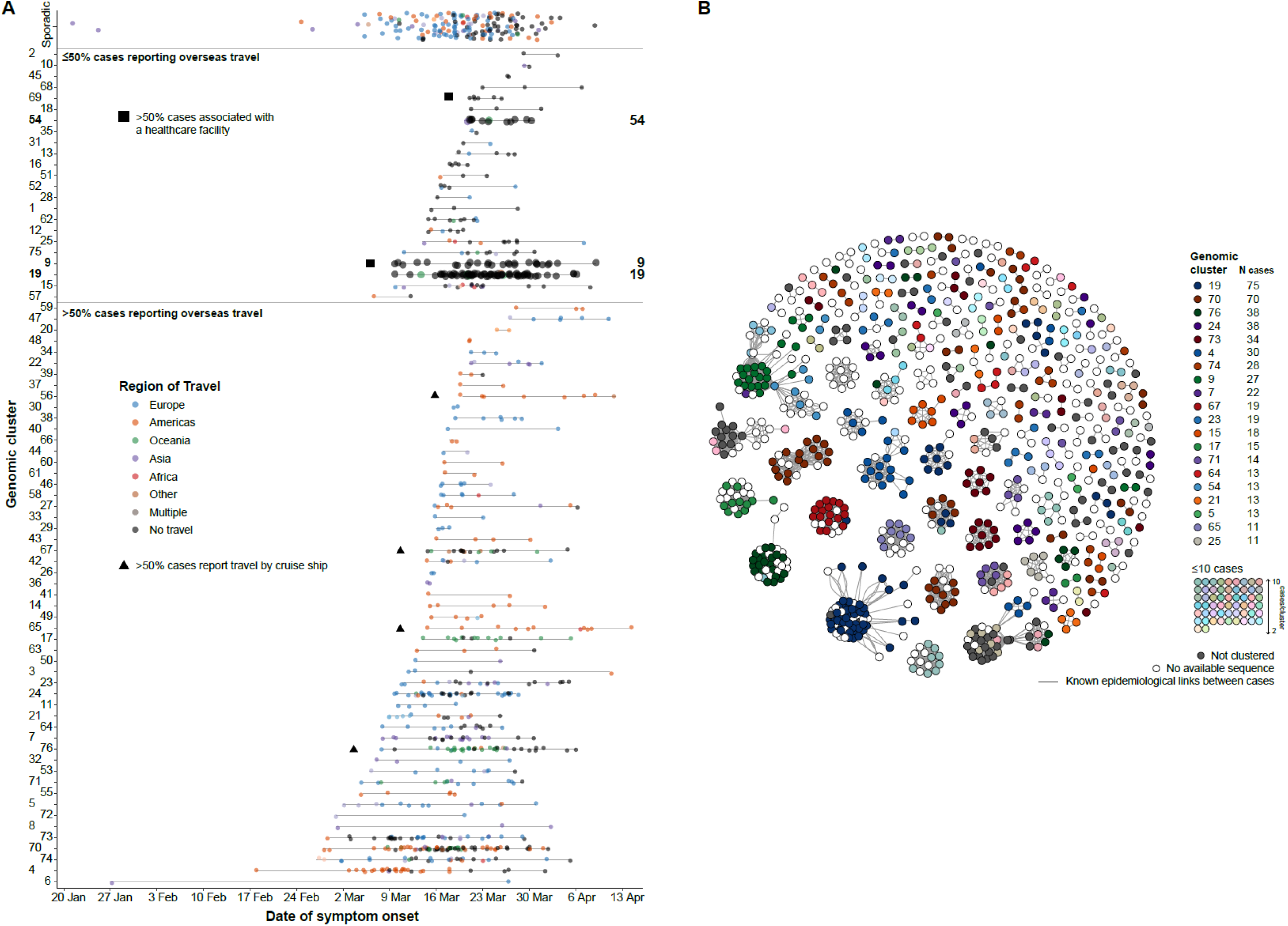
Combined genomic and epidemiologic data for Victorian SARS-CoV-2 cases. **A. Timeline and key epidemiological features of Victorian SARS-CoV-2 genomic clusters**. Each case in a genomic cluster is represented by a dot, colored by location of travel. Cases are plotted by onset date on the X-axis and genomic cluster on the Y-axis. Genomic clusters discussed in the text are enlarged and marked with their cluster number to the right. **B. Network analysis demonstrating concordance between genomic and epidemiological clustering amongst Victorian COVID-19 cases with known epidemiological links**. Each filled dot (node) represents a Victorian COVID-19 case with a documented epidemiological link to another Victorian case. Edges (links) between nodes (cases) represent each epidemiological link. Cases are placed closer to each other within the network as the density (number) of linkages between them increases, with cases in the same epidemiological cluster forming a spatially distinct group. Cases are colored by genomic cluster; cases where a sequence was not included primary analysis are colored white.

Of the 81/134 sequenced cases (61%) with an epidemiologically unknown source of acquisition, 71 (88%) were identified within 24 genomic clusters, providing insight into potential sources of acquisition for these epidemiologically undefined cases. This information was provided to the genomics response team to inform public health investigation in these cases.

### Investigation of transmission clusters of public health importance

Several genomic clusters were investigated further due to their size and high-risk settings (Figure 3). For example, 48/51 (94%) cases in four epidemiological clusters associated with social venues in a specific area of metropolitan Melbourne were found within genomic cluster 19 (total 75 cases), providing genomic evidence for community transmission. Importantly, 16/75 cases (21%) in this cluster had no known epidemiological source of infection, and 7/75 cases (9%) were associated with an epidemiologically unlinked health service.

Genomic analysis was also used to exclude a putative transmission network involving four health services, epidemiologically linked by common healthcare workers or healthcare exposure. Preliminary epidemiological analysis suggested this network comprised 54 cases; however, genomic investigations revealed the cases fell within at least four distinct genomic clusters (clusters 9, 69, 54, and 27, Figure 3). On further investigation, three cases from two health services found in Cluster 54 (total 15 cases) were determined to have attended the same social event, excluding healthcare associated transmission in one health service. Among our dataset, 4/76 (5%) genomic clusters had >50% of cases associated with a cruise ship (Figure 3). 17/74 cases (23%) in these clusters had no history of overseas travel, indicating limited onwards transmission.

### Genomic assessment of intra-patient diversity

Sixty-three cases had more than one sample sequenced over the study period (median 2 sequences per case, range 2-5), with a median of 10 days between first and last sample (IQR 5-13 days) (Supplementary Table S2 and Supplementary Figure S4). The median intra-patient pairwise SNP distance was 0 (range 0-18 SNPs), compared to a median inter-patient pairwise SNP distance of 11 SNPs (range 0-27 SNPs). Three intra-patient pairs were outliers, having pairwise SNP distances of 7, 9 and 18 SNPs. On manual inspection, at least one sequence from each pair was found to have more ambiguous or missing base calls than the rest of the dataset, potentially contributing to the high number of intra-patient SNPs. In order to further assess reproducibility of SARS-CoV-2 sequencing, duplicate sequencing of the same clinical sample was also performed across different sequencing runs for ten samples, with zero SNP differences detected between consensus sequences.

### Genomic inferences of the effect of population-level public health interventions on SARS-CoV-2 basic reproductive number

Bayesian phylodynamic analyses estimated the time to the most recent ancestor of the 903 samples in late December 2019 (95% credible interval, CI: 18th December to 30th December), with an evolutionary rate of 1.1×10^−3^ substitutions/site/year (8.7×10^−3^ − 1.28×10^−3^). The birth-death skyline model suggested a considerable change in *R_e_* around 27th March (CI: 23rd – 31st March). Prior to 27^th^ March, the estimated *R_e_* was 1.63 (CI: 1.45 – 1.8) with a subsequent decrease to 0.48 (CI: 0.27 – 0.69) after this time (Figure 4). Our estimated *R_e_* prior to the 27th March implied an epidemic doubling time of 11 days (CI: 8.3 – 14.4 days) The sampling proportion parameter (the probability of successfully sequencing an infected case) after the identification of the first case in Victoria was estimated at 0.88 (CI: 0.7 – 1.0), consistent with intensive sequencing efforts in Victoria, and in accordance with the proportion of samples obtained for sequencing from cases in Victoria (1,075/1,333; 80.6%).

**Figure 4:**
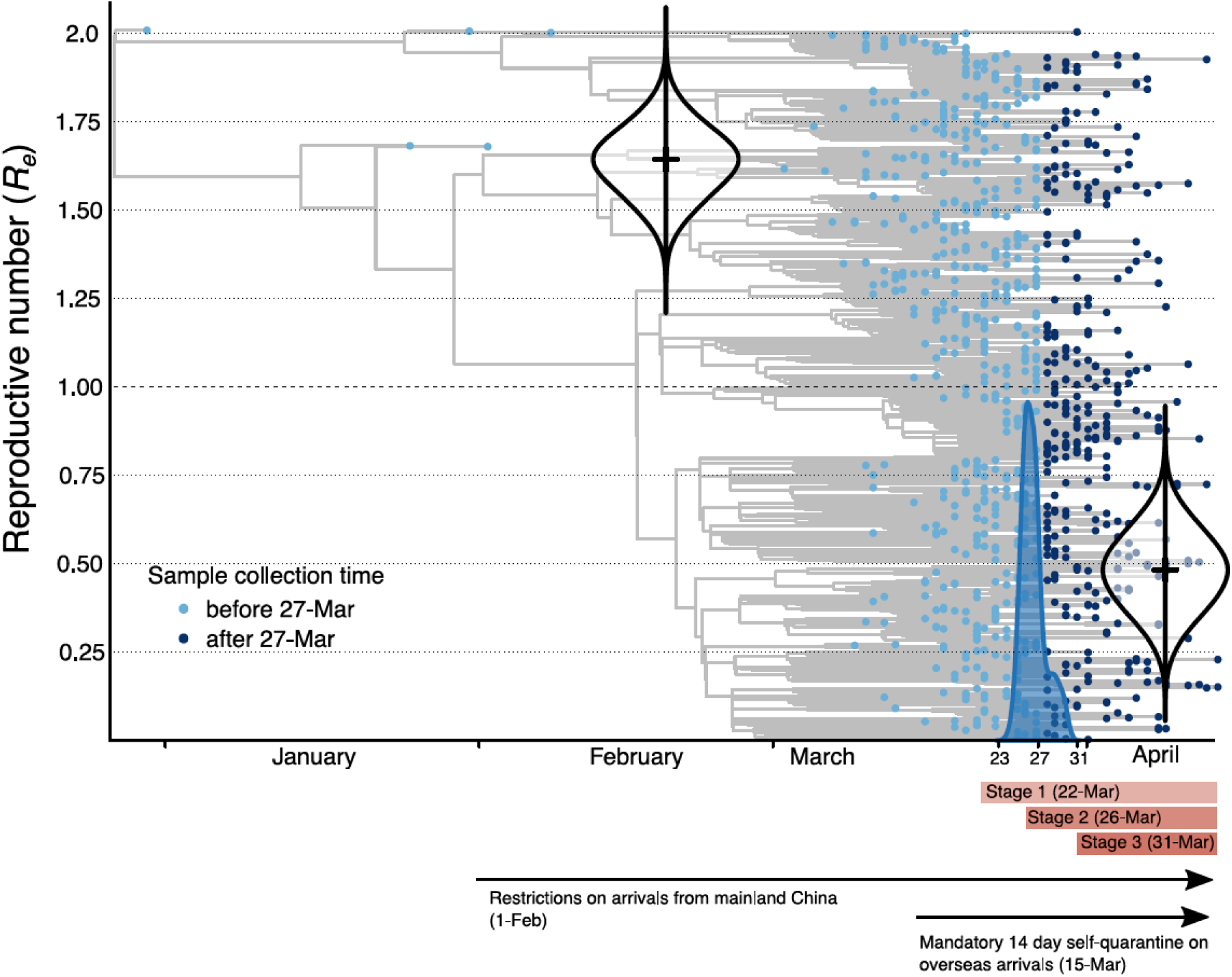
Phylodynamic estimates of the reproductive number (*R_e_*) A birth-death skyline model was fit, where *R_e_* is allowed to change at a single time point determined by the data. The x-axis represents time, from the molecular estimate of the origin of the sampled diversity, around late December 2019 (95% credible interval, CI: 18 December to 30 December) to the date of the most recently collected genome in 13 April. The blue shows the posterior distribution of the timing of the most significant change in *R_e_*, around 27 March (CI: 23 – 31 March). The y-axis represents *R_e_* and the violin plots show the posterior distribution of this parameter before and after the change in *R_e_*, with a mean of 1.63 (CI: 1.45 – 1.80) and 0.48 (CI:0.27 – 0.69), respectively. The phylogenetic tree in the background is a maximum clade credibility tree with the tips colored according to whether they were sampled before or after March 27^th^.

## DISCUSSION

We provide a detailed picture of the emergence and limited onward spread of SARS-CoV-2 in Australia, representing the largest genomic epidemiological study of SARS-CoV-2 to date. The sheer scale and rapidity of the COVID-19 pandemic has necessitated swift and unprecedented public health responses, and the high proportion of cases with associated sequence data in our study provides unique genomic insights into the effects of public health interventions on the spread of SARS-CoV-2. The genomic clusters in our dataset reflect some of the key public health and epidemiological themes that have emerged globally for COVID-19.

First, consistent with epidemiological data, 76% of genomic clusters in our setting had at least 50% of cases associated with international travel, although in most instances, there was only limited onward transmission. The changing origin of travel-associated clusters in our dataset (Asia, Europe, North America) is in keeping with the temporal emergence of these areas as global ‘hot-spots’ for COVID-19, and in keeping with sequential international travel restrictions declared by the Australian Government^18^. Of note, 22% of travel-associated cases were ‘sporadic’ (i.e. not in a genomic cluster), providing genomic evidence for the positive effects of widespread public health messaging and self-isolation requirements for returning travelers. Moreover, we identified four genomic clusters that were associated with cruise ship passengers either returning to or disembarking in Melbourne. Throughout the global COVID-19 pandemic, cruise ships have been identified as ‘amplification vessels’ for COVID-19, with onward seeding into ports.^19^ In Victoria, returning cruise ship passengers were quarantined on arrival, and our genomic data suggest only minimal onward transmission of infection from cruise ship passengers, supported by limited numbers of non-travel associated cases in these cluster, highlighting the effectiveness of local containment measures for this high-risk population group.

Second, consistent with the impact of COVID-19 in healthcare facilities in other settings, we identified a large genomic cluster of SARS-CoV-2 in a healthcare facility in Melbourne, with cases identified in patients and staff. Although genomics has been used extensively for infection control purposes in other pathogens, our data highlight the utility of genomics for SARS-CoV-2 infection control, with potential applications in monitoring the effectiveness of local policies for identifying high-risk patients, and in assessing the effectiveness of personal protective equipment (PPE). Applying genomics in healthcare settings is particularly important in the context of high reported rates of nosocomial acquisition of COVID-19 in other settings, with associated fatalities.^20,21^

Third, prior to the implementation of enhanced (stage 3) restrictions in Victoria, we identified a large genomic cluster (the largest in our dataset, comprising 75 cases) associated with several social venues in metropolitan Melbourne. This finding demonstrates the propensity for chains of SARS-CoV-2 transmission throughout urban areas associated with leisure activities and provides additional justification for the unprecedented population-level social restrictions in our setting. Further genomic support for the effectiveness of social restrictions is provided by our phylodynamic analysis, which demonstrates a decrease in *R_e_*, after the introduction of stage 3 restrictions (including mandatory quarantine in hotels for overseas returnees), from 1.63 to 0.48. The reduction in *R_e_* supports a decrease in disease incidence after the introduction of social restrictions, broadly in keeping with recent epidemiological modelling, suggesting a decrease in *R_e_* in Victoria around mid-March ^17^. The differences between epidemiological and genomic modelling may be due to differences in underlying models and the expected lag between demographic processes and their effect on molecular variation, even for rapidly evolving pathogens^22^.

A major strength of our study is that we were able to sequence samples from approximately 80% of all cases in Victoria, facilitated by the centralized nature of public health laboratory services in our setting. The high proportion of sequenced cases allowed us to address very specific queries from a public health perspective (e.g., whether case X belongs to cluster Y), enabling enhanced contact tracing when there was uncertainty around epidemiological information. Key to this effort was high-throughput sequencing using an amplicon-based approach, which allowed us to process a large number of samples in a short period of time. Stringent QC to ensure only high-quality consensus sequences entered the final alignment was particularly important when considering the minimal diversity in SARS-CoV-2 sequence data used to infer genomic clusters.^23,24^ While use of a predefined Ct value to select samples for SARS-CoV-2 genomic sequencing could be considered,^25^ our use of QC parameters, rather than a Ct value, enabled the inclusion of additional samples for genomic analysis, with samples with Ct values >40 being successfully included. Of further note was our assessment of intra-patient diversity, representing the largest analysis of intra-patient SARS-CoV-2 diversity to date. Our observation of minimal intra-patient SARS-CoV-2 diversity is in keeping with other recent findings,^26^ and provides additional evidence for the reproducibility of our sequencing and analysis.

In summary, we provide detailed genomic insights into the emergence and spread of SARS-CoV-2 in Australia and highlight the effect of public health interventions on the transmission of SARS-CoV-2. Through a combination of rapid public health responses, extensive diagnostic testing and collective social responsibility, Australia has successfully navigated the first wave of the COVID-19 pandemic. As social restrictions inevitably ease, the role of genomics will become increasingly important to rapidly identify and ‘stamp out’ possible transmission chains. Our data provide a framework for the future application of genomics in the response to COVID-19.

## ACKNOWLEDGEMENTS

We thank the public health, clinical, and microbiology staff across Victoria who have been involved in the testing, clinical care and public health responses to COVID-19. The data collected through public health officers and microbiology laboratories are critical for public health genomics investigations. We also thank Nick Loman, Jonathon Jacobs, Duncan Maccannell and Karthik Gangavarapu for bioinformatics advice, and Josh Quick, George Taiaroa and Sara Zufan for assistance with obtaining ARTIC primers early in the pandemic.

## AUTHOR CONTRIBUTIONS

B.H., D.W. and T.S. conceived of and designed the study. L.C, M.S., S.B., T.H., M.E., S.D., J.D., M.C., B.S., A.vD., and C.A. collected and generated data. T.S., C.L., N.S., S.D., A.G., K.H., T.S., C.A., D.W., and B.H. analysed the data. T.S., C.L., N.S., D.W., and B.H. wrote the manuscript. All authors reviewed and revised the manuscript.

## REFERENCES

1. Zhou P, Yang X-L, Wang X-G, et al. A pneumonia outbreak associated with a new coronavirus of probable bat origin. Nature 2020;579:270–3.

2. Zhu N, Zhang D, Wang W, et al. A novel coronavirus from patients with pneumonia in China, 2019. NEJM 2020; 382: 727–733.

3. Corman VM, Landt O, Kaiser M, et al. Detection of 2019 novel coronavirus (2019-nCoV) by real-time RT-PCR. Eurosurveillance 2020;25(3):pii=2000045.

4. Patel R, Babady E, Theel ES, et al. Report from the American Society for Microbiology COVID-19 International Summit, 23 March 2020: Value of Diagnostic Testing for SARS–CoV-2/COVID-19. mBio 2020; 11:e00722–20.

5. Forster P, Forster L, Renfrew C, Forster MJPotNAoS. Phylogenetic network analysis of SARS-CoV-2 genomes. 2020;117:9241–3.

6. Stefanelli P, Faggioni G, Presti AL, et al. Whole genome and phylogenetic analysis of two SARS-CoV-2 strains isolated in Italy in January and February 2020: additional clues on multiple introductions and further circulation in Europe. Eurosurveillance 2020; 25(3):pii=2000305.

7. Chan JF-W, Yuan S, Kok K-H, et al. A familial cluster of pneumonia associated with the 2019 novel coronavirus indicating person-to-person transmission: a study of a family cluster. Lancet 2020;395:514–23.

8. Sansone M, Andersson M, Gustavsson L, Andersson L-M, Nordén R, Westin JJCID. Extensive hospital in-ward clustering revealed by molecular characterization of influenza A virus infection. Clin Infect Dis 2020; ciaa108.

9. Poon AF, Gustafson R, Daly P, et al. Near real-time monitoring of HIV transmission hotspots from routine HIV genotyping: an implementation case study. Lancet HIV 2016; 3:e231-e8.

10. Peters PJ, Pontones P, Hoover KW, et al. HIV infection linked to injection use of oxymorphone in Indiana, 2014–2015. NEJM 2016;375:229–39.

11. Caly L, Druce J, Roberts J, et al. Isolation and rapid sharing of the 2019 novel coronavirus (SARS-CoV-2) from the first patient diagnosed with COVID-19 in Australia. Med J Aust 2020; mja20.00186(2).

12. ARTIC-nCoV2019 primer schemes (Github). 2020. at https://github.com/artic-network/artic-ncov2019/tree/master/primer_schemes/nCoV-2019/V3.

13. nCoV-2019 sequencing protocol. 2020. at https://dx.doi.org/10.17504/protocols.io.bbmuik6w.

14. Ragonnet-Cronin M, Hodcroft E, Hué S, et al. Automated analysis of phylogenetic clusters. BMC Bioinformatics 2013; 14: 317.

15. Rambaut A, Holmes EC, Hill V, et al. A dynamic nomenclature proposal for SARS-CoV-2 to assist genomic epidemiology. bioRxiv 2020, https://doi.org/10.1101/2020.04.17.046086.

16. Bouckaert R, Vaughan TG, Barido-Sottani J, et al. BEAST 2.5: An advanced software platform for Bayesian evolutionary analysis. J PLoS Comp Biol 2019;15:e1006650.

17. Theoretical modelling to inform Victoria’s response to coronavirus (COVID-19). 2020. at https://www.dhhs.vic.gov.au/theoretical-modelling-inform-victorias-response-coronavirus-covid-19.

18. COVID-19 advice for travellers. 2020. at https://www.smartraveller.gov.au/COVID-19-australian-travellers.

19. Moriarty LF, Plucinski MM, Marston BJ, et al. Public Health Responses to COVID-19 Outbreaks on Cruise Ships - Worldwide, February-March 2020. MMWR 2020;69:347–52.

20. Zhan M, Qin Y, Xue X, Zhu S. Death from Covid-19 of 23 Health Care Workers in China. NEJM 2020; doi: 10.1056/NEJMc2005696.

21. Wu Z, McGoogan JM. Characteristics of and Important Lessons From the Coronavirus Disease 2019 (COVID-19) Outbreak in China: Summary of a Report of 72314 Cases From the Chinese Center for Disease Control and Prevention. JAMA 2020; 323(13):1239–1242.

22. du Plessis L, Stadler T. Getting to the root of epidemic spread with phylodynamic analysis of genomic data. Trends Microbiol 2015;23:383–6.

23. Lu R, Zhao X, Li J, et al. Genomic characterisation and epidemiology of 2019 novel coronavirus: implications for virus origins and receptor binding. Lancet 2020;395:565–74.

24. Pung R, Chiew CJ, Young BE, et al. Investigation of three clusters of COVID-19 in Singapore: implications for surveillance and response measures. Lancet 2020.

25. Rockett RJ, Arnott A, Lam C, et al. Revealing COVID-19 transmission by SARS-CoV-2 genome sequencing and agent-based modelling. bioRxiv2020.04.19.048751; https://doi.org/10.1101/2020.04.19.048751.

26. Wolfel R, Corman VM, Guggemos W, et al. Virological assessment of hospitalized patients with COVID-2019. Nature 2020; 1–10.

